# Predicting Total Knee Replacement in Knee Osteoarthritis Using a Machine-Learning–Guided Approach in patients of the Osteoarthritis Initiative (OAI)

**DOI:** 10.1101/2025.10.28.25338966

**Authors:** Francisco J. Blanco, Natividad Oreiro, Jorge Vázquez-García, Antonio Morano-Torres, Vanesa Balboa-Barreiro, Isabel Rodríguez-Valle, Sara Relaño, Nicola Veronese, María C. de Andrés, Ignacio Rego-Pérez

## Abstract

**Objective:** To develop a pragmatic model to predict total knee replacement (TKR) in knee osteoarthritis (OA) using non-imaging clinical, genetic, and lifestyle data with machine-learning (ML)–guided feature selection.

**Methods:** We analyzed 3,790 Osteoarthritis Initiative (OAI) participants. Nested ML feature selection on the training set identified 15 informative variables. Classifiers were benchmarked, then a multivariable logistic regression was fit on the full cohort. Performance was summarized by discrimination (AUC with 95% CI) and calibration (Brier score). To assess the incremental value of genetics, we refit an otherwise identical Clinical model excluding the polygenic risk score (PRS) and compared specificity at fixed sensitivities using Bonferroni-adjusted McNemar tests. A pre-specified analysis examined performance by baseline Kellgren–Lawrence (KL) grade (KL 0–1 vs KL ≥2).

**Results:** On the test set, classifier AUCs ranged 0.716–0.748, with Elastic Net and XGBoost performing best. The final logistic model fit on the full cohort achieved AUC 0.765 (95% CI 0.736–0.793) with acceptable calibration (Brier 0.097). Performance remained robust by disease stage, with higher discrimination in pre-radiographic knees (KL 0–1: AUC 0.827) and moderate discrimination in KL ≥2 (AUC 0.720); decile plots indicated broadly aligned observed vs predicted risks. PRS added modest, statistically significant gains in specificity at several fixed sensitivities without materially changing AUC.

**Conclusions:** We present a pragmatic, non-imaging, ML-informed model that predicts TKR with clinically acceptable discrimination and calibration using routinely collected data. This framework provides a practical basis for individualized risk stratification and decision support without reliance on imaging.

**What is already known on this topic:** Risk of total knee replacement (TKR) in knee osteoarthritis (OA) is multifactorial and many existing models depend on imaging markers such as Kellgren–Lawrence grade or MRI findings. Established non-imaging predictors include symptoms and function (WOMAC), age, BMI, knee alignment or prior injury. Genetic scores have been explored in OA but, to date, have shown limited standalone utility compared with routine clinical factors.

**What this study adds:** This study presents a clinic-friendly, non-imaging prediction model guided by a transparent ML pipeline—nested random-forest feature selection with in-fold preprocessing and SMOTE, repeated cross-validation, and SHAP-based interpretation—that achieves acceptable discrimination and calibration in the OAI cohort. It reinforces the relevance of routine clinical factors, identifies an inverse association between Mediterranean-diet adherence and TKR risk, and evaluates the incremental—though limited—contribution of genetic risk via a polygenic risk score (PRS), with a signal that persists in pre-radiographic knees despite few events.

**How this study might affect research, practice or policy:** The model offers a practical pathway for risk stratification where imaging is unavailable or costly, supporting shared decision-making and prioritization of follow-up. It encourages precision-medicine workflows that integrate clinical and genetic information cautiously and transparently, and it sets clear directions for future work: external validation across settings, assessment in early-stage OA populations, and refinement of genetic predictors before any policy or guideline incorporation.

## INTRODUCTION

Osteoarthritis (OA) is a chronic musculoskeletal disease of polygenic and heterogeneous nature, primarily affecting movable joints. Globally, over 500 million people have OA [1], with the knee being the most commonly affected joint [2–4]. Symptoms include activity-related pain, often worsened by weight-bearing and relieved by rest. While age is a major risk factor, other contributors include genetics, joint trauma, malalignment, muscle weakness, joint overload, and female sex [5]. Modifiable risk factors such as mechanical stress, obesity, and overweight are especially relevant [6].

OA is traditionally considered slowly progressive, with many patients worsening over decades. Around 20% reach end-stage disease requiring joint replacement, a major surgery with notable morbidity and mortality [7]. Total knee replacement (TKR) should only be considered when other treatments fail [5]. However, TKR rates are rising—from 120 surgeries per 100,000 people in 2014 to 165 in 2022—and are expected to increase substantially with aging populations [8–10]. This trend poses a growing burden on healthcare systems. Additionally, up to 20% of TKR patients experience persistent postsurgical pain due to various surgical, prosthetic, or patient-specific causes [11].

Genetic heritability of knee OA is estimated at 45–60% [12]. With advances in genome-wide association studies (GWAS) [13, 14], polygenic risk scores (PRS) have emerged as tools with potential clinical value [15]. PRS condense genetic susceptibility to a trait by combining weighted risk alleles from GWAS summary statistics. Several studies have assessed PRS for TKR risk in European [16, 17] and Japanese cohorts [18] using one of the largest OA GWAS to date [13], though, to our knowledge, none of these studies used the Osteoarthritis Initiative (OAI) as the target cohort.

Developing predictive models to identify patients at high TKR risk is crucial for enabling lifestyle interventions and personalized care. Machine learning (ML) models are particularly suited to capture complex interactions among clinical, molecular, and genetic variables. In TKR prediction, they handle class imbalance and improve model accuracy while also aiding interpretation by selecting the most relevant predictors. This supports both risk prediction and data-driven preventive strategies. Application of ML methods is an emerging field in OA and particularly, in knee OA [19–21], including for TKR risk prediction [22–24]. Most of these rely on imaging data, such as X-rays or MRIs, which involve cost and, in the case of X-rays, repeated radiation exposure.

This study sought to build and internally evaluate a non-imaging, ML-guided clinical model for predicting TKR in the OAI cohort by quantifying discrimination and calibration after ML-based feature selection, benchmarking against alternative classifiers, performing pre-specified KL-stratified analyses, and, secondarily, exploring the incremental contribution of genetic information via a PRS.

## METHODS

An overview of the study methodology is shown in **Figure 1**. All analyses were conducted using R v4.4.1.

**Figure 1.**
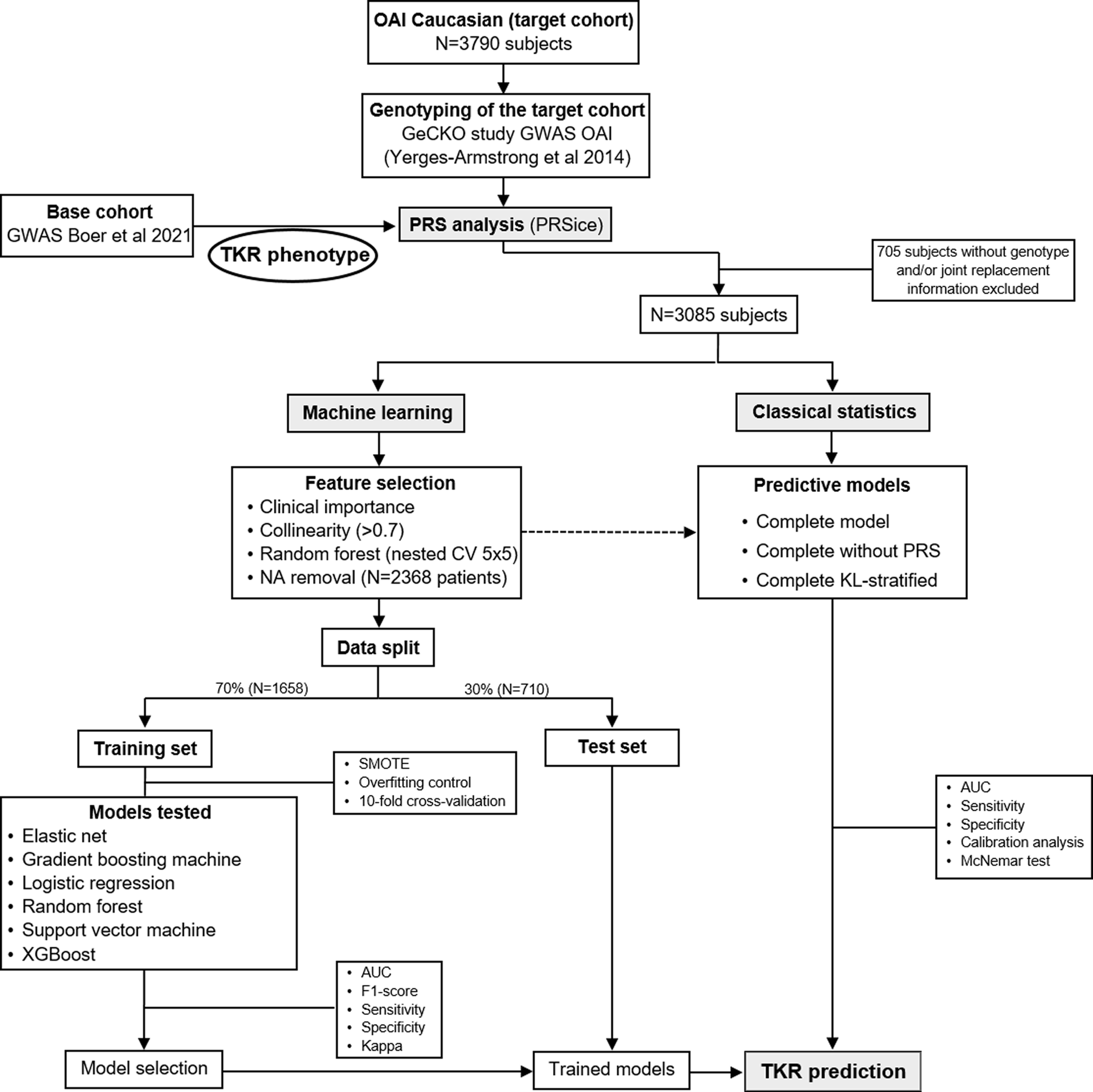
Scheme and flow chart of the methodology used in this study. OAI: Osteoarthritis Initiative; TKR: total knee replacement; PRS: polygenic risk score; SMOTE: synthetic minority oversampling technique; GeCKO: Genetic components of knee Osteoarthritis.

### Cohort description

We used data from the Osteoarthritis Initiative (OAI), a well-characterized prospective cohort of knee OA patients with publicly available data and biospecimens (https://nda.nih.gov/oai) [25]. Participants were followed annually for 108 months to track symptom progression, disability, structural changes, and biomarker levels. All study procedures complied with the Declaration of Helsinki, and participants gave informed consent.

### Machine learning approach

All ML analyses and their reporting followed the Transparent Reporting of a multivariable prediction model for Individual Prognosis Or Diagnosis with Artificial Intelligence (TRIPOD+AI) guidelines [26]. A completed checklist is provided in the Supplementary Material. We used ML to identify (non-imaging) predictors and construct an optimized model for TKR risk.

### Data source and pre-processing

We obtained all non-imaging variables from the OAI and randomly split the data into a training set (70%) and a test set (30%). First, we retained only expert-curated and literature-supported predictors [27], then removed highly collinear features (Pearson correlation >0.7). To reduce dimensionality without information leakage, we applied a nested Random Forest (rf) feature-selection framework (5 outer folds x 5 inner folds) on the training data. In each outer fold, the top-15 variables by averaged importance were recorded, and the final predictor set was the consensus of variables appearing in ≥3/5 outer folds. Class imbalance during feature selection was handled via class weighting, and all preprocessing steps (including centering/scaling) were fit within training folds and then applied to the corresponding validation/test partitions.

### Additional data source used in the process

We additionally considered three variables as candidate predictors: (i) adherence to the Mediterranean diet (MD), ii) a published genetic interaction between the GG genotype of SNP rs12107036 in TP63 and mtDNA haplogroup UK [28], and iii) genetic OAI data used to construct a PRS for TKR risk. All three were added via expert curation and processed identically to the other candidates within the nested rf feature-selection framework, without being forced into the models.

### Mediterranean diet (MD)

MD adherence, provided by Veronese et al. [29], was assessed using the alternate Mediterranean diet (aMED) score [30]. This score is based on a food frequency questionnaire, which was recorded during the baseline OAI visit. The aMED takes into consideration several foods commonly consumed within the MD, including: cereals, potatoes, fruits, vegetables, legumes, fish, red meat, poultry, full fat dairy products, olive oil and consumption of alcoholic beverages. The intake of each food was categorized on the basis of monthly, weekly, or daily consumption, as appropriate, ranging from a score of 0 (less adherent) to 5 (better adherence); the total score ranges from 0 to 55, with higher values indicating higher adherence to a MD. Since there are no agreed cut-off scores for higher aMED adherence, the population was divided into quartiles: aMED <25, 26-28, 29-32, and ≥33.

### Genetic interaction

An interaction between the TP63 rs12107036 GG genotype and mtDNA haplogroup UK has been reported as a significant risk factor for rapidly progressive knee OA in OAI participants [28]. Accordingly, we included this interaction as a candidate predictor of TKR risk.

### PRS construction

Genotype data from 4,219 OAI participants were obtained from dbGAP (phs000955.v1.p1), as part of the genetic components of knee osteoarthritis (GeCKO) study [31], on the Illumina HumanOmni2.5 BeadChip. Variant-level QC excluded markers failing call rate, duplication or monomorphism, with MAF < 0.05, or Hardy–Weinberg equilibrium p-value < 1×10⁻D. Sample-level QC removed individuals with >3% missing genotypes, outlier heterozygosity (±4 SD), duplicates/relateds, and ancestry outliers (>8 SD from the European 1000 Genomes PCA cluster). Post-QC VCFs were imputed on the Michigan Imputation Server (Minimac4), retaining variants with Rsq ≥ 0.8 for high-confidence analyses (≈8 million variants).

To verify allele alignment, population structure control and genomic inflation, we ran a screening association scan using rvtests [32] with the score model, including as covariates the individual’s age, sex, body mass index (BMI), as well as principal components (PCs) derived from a pruned hard-call SNP set under stricter missingness/relatedness thresholds. This step was not intended for SNP discovery and no SNP-level results are reported. The final dataset included 3,085 individuals.

PRS was derived from published OA GWAS summary statistics [13]. We selected 25 significantly associated with TKR risk (p-value < 1.3e^-8^) as the base set of instruments and computed a weighted score in PRSice using the reported log(OR) for the effect alleles. Effect/other alleles were aligned to the base GWAS; palindromic SNPs at ambiguous MAF were excluded or resolved using reference frequencies. Using OAI imputed data as the target, clumping in PRSice (r² = 0.1) retained 12 independent SNPs after LD pruning, with age, sex, BMI and the aforementioned PCs entered as covariates (**Supplementary Table 1**). The PRS was standardized (z-score) and categorized by quintiles into low (Q1, 0–20%), medium (Q2–Q4, 21–80%) and high (Q5, 81–100%) risk groups [16].

### Construction of ML models

We trained six supervised models using caret: Random Forest (rf), XGBoost (xgbTree), Logistic Regression (glm), Gradient Boosting Machine (gbm), Elastic Net (glmnet), and Support Vector Machine (svmRadial). Hyperparameters were tuned via 10-fold cross-validation with centering/scaling and Synthetic Minority Over-sampling Technique (SMOTE; k = 5) applied within each fold. Using the 15 predictors selected in the nested feature-selection step, we then ran a repeated 5-fold cross-validation (5 repeats) with the same in-fold preprocessing and SMOTE to obtain stable performance estimates; we report mean ± SD AUC, considering AUC > 0.70 as clinically acceptable [33]. Finally, each model was refit on the full training data and evaluated once on the independent test set, reporting unbiased AUC, sensitivity, specificity, Kappa, and F1-score. The operating threshold was chosen by Youden’s index on the training set and then applied to the test set.

In addition, we used SHAP (SHapley Additive exPlanations) to quantify the contribution of each predictor to individual model outputs. This approach, assigns an importance value to each feature based on its impact on individual predictions. Violin plots (summary plots) showed the distribution of feature importance across samples.

### Statistical analyses

After the subsequent process of ML to select the most informative non-imaging predictors, a multivariable logistic regression model was developed on the entire cohort. Predictive performance of the model was summarized by discrimination (area under the ROC curve, AUC, with 95% confidence interval (CI)) and calibration (Brier score and decile-based calibration plots with 95% CIs).

As a secondary analysis, we examined whether genetic information (in terms of PRS) adds value beyond clinical predictors. To do this, we fitted a Clinical model identical to the Complete model but excluding the PRS. The AUC values of these two models were compared using the DeLonǵs test. Calibration was also evaluated via the Brier score and decile-based calibration plots with 95% CI. In addition, McNemar tests with Bonferroni correction evaluated specificity differences between models at fixed sensitivity levels (0.400–0.900). Confusion matrices were also constructed.

### KL-stratified analysis

To explore stage-related performance, the Complete model was re-estimated within baseline KL 0–1 and KL ≥2 strata. Within each stratum, we reported AUC, Brier score, decile-based calibration plots, and coefficient estimates for all predictors, following the same procedures as in the primary analysis.

## RESULTS

### Machine learning approach. Data distribution and model performance

Fifteen final predictors were selected (**Table 1**). After removing NAs, the training set included 1,658 individuals (737 males, 921 females), with 202 (12.2%) undergoing TKR. The test set included 710 individuals (314 males, 396 females), of whom 89 (12.5%) had TKR. Feature distributions did not significantly differ between sets.

**Table 1.** Feature selection process in training and test sets.

| Feature | Training set<br>(N=1658) | Test set<br>(N=710) |
| --- | --- | --- |
| BMI (Kg/m<sup>2</sup>) | 28.11±4.59 | 28.10±4.54 |
| Age at baseline (years) | 61.32±8.99 | 61.85±9.19 |
| Sex: |  |  |
| Male | 737 (44.50) | 314 (44.20) |
| Female | 921 (55.50) | 396 (55.80) |
| Standardized PRS (per SD) | 0.01±1.01 | -0.03±0.97 |
| aMED score | 28.29±5.00 | 28.76±4.79 |
| Systolic pressure (mm Hg) | 122.61±15.70 | 123.7±15.26 |
| Diastolic pressure (mm Hg) | 74.64±9.52 | 74.59±10.35 |
| NSAID exposure: |  |  |
| No-exposure | 643 (38.80) | 277 (39.0) |
| Exposure | 1015 (61.20) | 433 (61.0) |
| Knee alignment: |  |  |
| Normal | 342 (20.60) | 133 (18.70) |
| Varus | 493 (29.70) | 219 (30.80) |
| Valgus | 823 (49.60) | 358 (50.40) |
| TP63-GGxUk<sup>#</sup> |  |  |
| GGxUk | 99 (6.0) | 48 (6.80) |
| GGxno-Uk | 360 (21.70) | 144 (20.30) |
| No-GGxUk | 283 (17.10) | 126 (17.70) |
| No-GGxNo-Uk | 916 (55.20) | 392 (55.20) |
| Left knee injury: |  |  |
| Yes | 458 (27.60) | 201 (28.30) |
| No | 1200 (72.40) | 509 (71.70) |
| Right knee injury: |  |  |
| Yes | 527 (31.80) | 196 (27.60) |
| No | 1131 (68.20) | 514 (72.40) |
| WOMAC disability | 11.98±14.82 | 13.22±16.47 |
| WOMAC pain | 3.58±4.16 | 4.00±4.82 |
| Vitamin C suppl (mg/day)* | 299.81±458.32 | 328.63±502.63 |
| Total knee replacement: |  |  |
| Yes | 202 (12.20) | 89 (12.50) |
| No | 1456 (87.80) | 621 (87.50) |

Values are mean±standard deviation or number of patients with percentage in parentheses; BMI: body mass index; PRS: polygenic risk score; NSAID: non-steroid anti-inflammatory drugs; WOMAC: Western Ontario and McMaster Universities Osteoarthritis Index; aMED: adherence to Mediterranean diet; (\*) GG genotype of single nucleotide polymorphism (SNP) rs12107036 at *TP63* and mitochondrial DNA haplogroup Uk; (\*) average daily nutrients from vitamin C supplements (in milligrams).

**Table 2** shows model performance. In the test set, AUC values ranged from 0.716 (Logistic Regression) to 0.748 (XGBoost). The highest F1-score was for XGBoost (0.378), followed by Elastic Net (0.376). Based on AUC and F1-score, Elastic Net (a regularized linear model) and XGBoost (a nonlinear model based on assemblies of decision trees) were the top-performing models. Confusion matrices are shown in **Figure 2a**.

**Figure 2.**
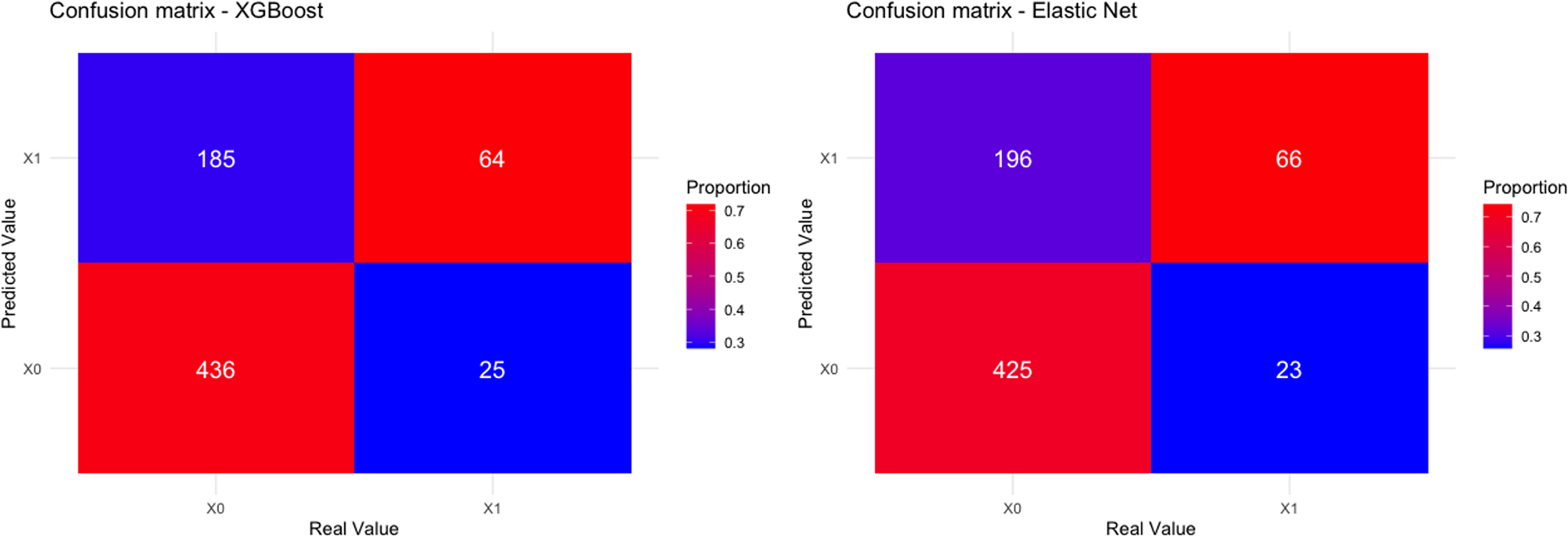
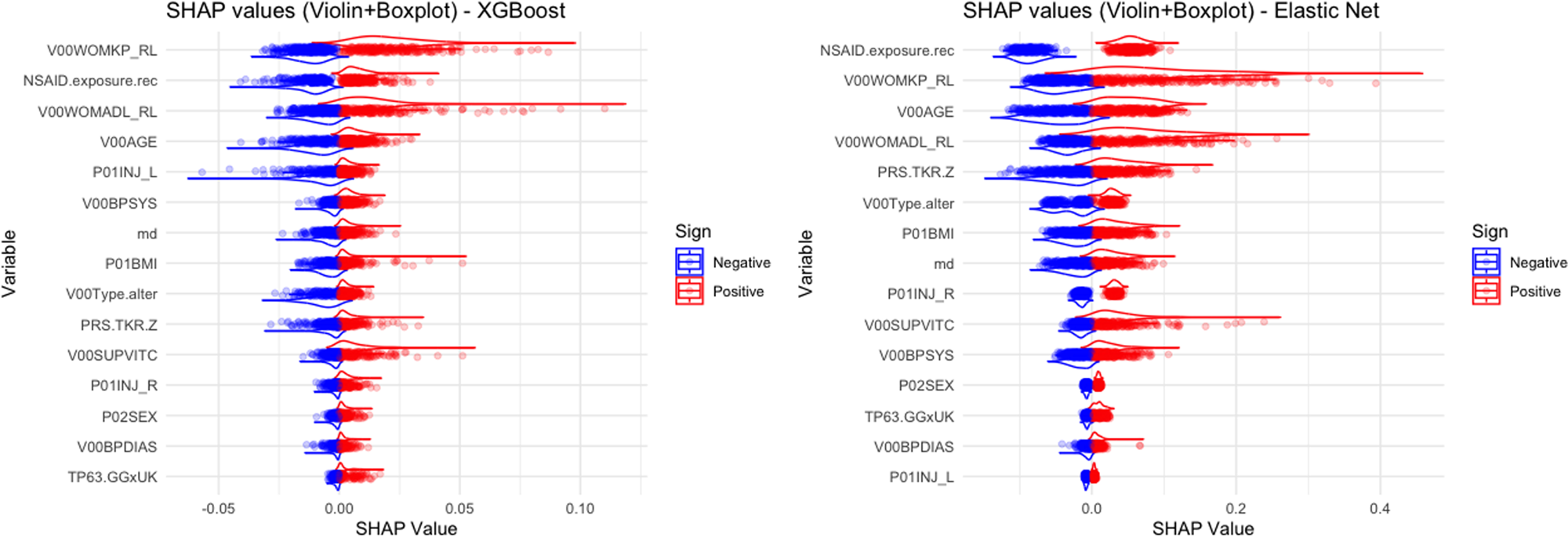
Confusion matrices and variable importance graphics. **A)** Confusion matrices of the XGBoost and Elastic net models. **B)** SHAP summary plot showing the impact of the most influential variables on the XGBoost and Elastic net model’s predictions. Each row represents a variable, with SHAP values on the x-axis indicating the magnitude and direction of its effect. Red denotes a positive contribution to the prediction, while blue indicates a negative one. Variables are ordered from top to bottom according to their mean absolute SHAP value. NSAID.exposure.rec: non-steroid anti-inflammatory drugs exposure; P01.INJ_R: previous injury in right knee; P01.INJ_L: previous injury in left knee; PRS.TKR.Z: standardized polygenic risk score; V00Type.alter: knee alignment (varum/valgus/normal); V00WOMKP_RL: WOMAC pain in right and/or left knee; P02SEX: sex; TP63.GGxUk: genetic interaction between SNP rs12107036 at *TP63* and mitochondrial DNA (mtDNA) haplogroup Uk; P01BMI: body mass index; V00AGE: age; md: adherence to Mediterranean diet; V00WOMADL_RL: WOMAC physical function; V00BPSYS: systolic pressure; V00BPDIAS; diastolic pressure; V00SUPVITC: Vitamin C supplementation.

**Table 2.** Metrics of the different machine learning models in the test set of the OAI.

| Model | AUC (95%CI) | F1-score | Sensitivity | Specificity | Kappa |
| --- | --- | --- | --- | --- | --- |
| Elastic net | 0.726 (0.670-0.782) | 0.376 | 0.741 | 0.684 | 0.232 |
| Gradient boosting machine (GBM) | 0.737 (0.684-0.791) | 0.358 | 0.618 | 0.738 | 0.219 |
| Logistic regression | 0.716 (0.659-0.774) | 0.364 | 0.719 | 0.680 | 0.217 |
| Random forest | 0.747 (0.698-0.798) | 0.229 | 0.168 | 0.956 | 0.162 |
| Support vector machine (SVM) | 0.726 (0.669-0.782) | 0.365 | 0.652 | 0.725 | 0.225 |
| XGBoost | 0.748 (0.696-0.800) | 0.379 | 0.719 | 0.702 | 0.238 |

AUC: Area under the curve; CI: confidence interval; OAI: Osteoarthritis initiative; GBM: gradient boosting machine; SVM: support vector machine

### Model predictors

The magnitude and direction of the effect of each variable on the prediction was determined by plotting the SHAP values on violin plots, where variables are ranked from top to bottom according to their overall importance, calculated as the mean absolute SHAP value across all observations (**Figure 2b**). In the case of XGBoost, visualization highlights that variables such as The Western Ontario and McMaster Universities Osteoarthritis Index (WOMAC) pain, non-steroid anti-inflammatory drugs (NSAIDs) exposure, WOMAC physical function, age and previous knee injury had the greatest influence on model output. For Elastic net, the most decisive features in this model was the NSAIDs exposure, followed by WOMAC pain, age, WOMAC physical function and standardized PRS.

### Final predictive model

We used a complete-case approach for the final logistic regression, including only participants with non-missing data for all model variables; no imputation was performed. Following ML-guided feature selection, the final Complete model was fitted in 2,414 participants and comprised 11 predictors (**Table 3**). For transparency, univariate associations for each predictor (ORs, 95% CIs, and p-values) are also reported: sex (OR=1.151; 95%CI=0.923– 1.436; p-value=0.211), age (OR=1.038; 95%CI=1.026–1.051; p-value <0.001), BMI (OR=1.074; 95%CI=1.050–1.098; p-value <0.001), GG genotype of SNP rs12107036 at *TP63* and mtDNA haplogroup Uk (GG.no-Uk vs GG.Uk: OR=0.551; 95%CI=0.352–0.862; p-value=0.009; no-GG.Uk vs GG.Uk: OR=0.509; 95%CI=0.319–0.812; p-value=0.005; no-GG.no-Uk vs GG.Uk: OR=0.624; 95%CI=0.418–0.932; p-value=0.021), standardized PRS (OR=1.198; 95%CI=1.069–1.344; p-value=0.002), systolic pressure (OR=1.696; 95%CI=1.360-2.114; p-value<0.001), NSAID exposure (OR=2.885; 95%CI=2.179–3.822; p-value<0.001), previous knee injury (OR=1.496; 95%CI=1.202–1.864; p-value<0.001), knee malalignment (varus vs normal: OR=1.491; 95%CI=1.067–2.084; p-value=0.019 and valgus vs normal: OR=1.429; 95%CI=1.045–1.956; p-value=0.026) WOMAC pain (OR=1.124; 95%CI=1.103–1.145; p-value<0.001) and aMED score (OR=0.974; 95%CI=0.953–0.996; p-value=0.021) (**Table 3**).

**Table 3.** Uni and multivariate regression analyses to predict the risk of TKR in 2,414 OAI patients.

|  | Univariate analysis |  | Multivariate analysis |  |
| --- | --- | --- | --- | --- |
|  | OR (95% CI) | p-value | OR (95% CI) | p-value |
| Sex (female) | 1.151 (0.923-1.436) | 0.211 | 1.090 (0.831-1.430) | 0.532 |
| Age at baseline (years) | 1.038 (1.026-1.051) | **<0.001\*** | 1.043 (1.027-1.059) | **<0.001<sup>#</sup>** |
| BMI (Kg/m<sup>2</sup>) | 1.074 (1.050-1.098) | **<0.001\*** | 1.039 (1.009-1.069) | **0.009<sup>#</sup>** |
| *TP63-GGxUK<sup>§</sup>* |  |  |  |  |
| GGxUk | 1 | - | 1 | - |
| GGxno-Uk | 0.551 (0.352-0.862) | **0.009\*** | 0.509 (0.300-0.863) | **0.012<sup>#</sup>** |
| No-GGxUk | 0.509 (0.319-0.812) | **0.005\*** | 0.534 (0.310-0.920) | **0.024<sup>#</sup>** |
| No-GGxNo-Uk | 0.624 (0.418-0.932) | **0.021\*** | 0.607 (0.380-0.970) | **0.037<sup>#</sup>** |
| Standardized PRS (per SD) | 1.198 (1.069-1.344) | **0.002\*** | 1.237 (1.086-1.410) | **0.001<sup>#</sup>** |
| PRS category: |  |  |  |  |
| Low-risk (Q1) | 1 | - |  |  |
| Medium-risk (Q2-Q4) | 1.338 (0.967-1.851) | 0.079 |  |  |
| High-risk (Q5) | 1.731 (1.195-2.507) | **0.004\*** |  |  |
| Systolic pressure (mm Hg) | 1.696 (1.360-2.114) | **<0.001\*** | 1.476 (1.128-1.929) | **0.004<sup>#</sup>** |
| NSAID exposure: |  |  |  |  |
| No-exposure | 1 | - | 1 | - |
| Exposure | 2.885 (2.179-3.822) | **<0.001\*** | 2.109 (1.544-2.881) | **<0.001<sup>#</sup>** |
| Previous knee injury |  |  |  |  |
| No-injury | 1 | - | 1 | - |
| Injury | 1.496 (1.202-1.864) | **<0.001\*** | 1.383 (1.060-1.805) | **0.017<sup>#</sup>** |
| Knee alignment: |  |  |  |  |
| Normal | 1 | - | 1 | - |
| Varus | 1.491 (1.067-2.084) | **0.019\*** | 1.570 (1.058-2.330) | **0.025<sup>#</sup>** |
| Valgus | 1.429 (1.045-1.956) | **0.026\*** | 1.452 (1.009-2.091) | **0.045<sup>#</sup>** |
| WOMAC pain | 1.124 (1.103-1.145) | **<0.001\*** | 1.123 (1.094-1.151) | **<0.001<sup>#</sup>** |
| aMED score | 0.974 (0.953-0.996) | **0.021\*** | 0.975 (0.949-1.002) | 0.069 |

TKR: total knee replacement; OAI: Osteoarthritis initiative; BMI: body mass index; PRS: polygenic risk score; SD: standard deviation; NSAID: non-steroid anti-inflammatory drug; WOMAC: Western Ontario and McMaster Universities Osteoarthritis Index; aMED: adherence to Mediterranean diet; OR: odds ratio; CI: confidence interval; Q1: quintile 1; (<sup>§</sup>) GG genotype of single nucleotide polymorphism (SNP) rs12107036 at *TP63* and mitochondrial DNA haplogroup Uk; (\*) significant after univariate analysis; (#) significant after multivariate analysis; significant p-values are **in bold**.

### Analysis of the incremental value of the PRS

Of the 3,790 OAI subjects of Caucasian ancestry with available genetic data, PRS was calculated for 3,085 participants with knee prosthesis information (Yes/No). Based on the PRS distribution, 608 (19.7%) were categorized as low-risk, 1,859 (60.3%) as medium-risk, and 618 (20%) as high-risk. **Table 4** summarizes sex, age, BMI, and knee replacement across PRS groups, with TKR frequency rising from 8.2% in low-risk to 10.7% in moderate-risk and 13.4% in high-risk (p-value=0.013). No significant interaction was found between PRS and sex or MD (data not shown).

**Table 4.** Baseline characteristics of the OAI population based on PRS.

|  | Low-risk PRS<br>(N=608) | Medium-risk PRS<br>(N=1859) | High-risk PRS<br>(N=618) | p-value |
| --- | --- | --- | --- | --- |
| Age at baseline (years) | 61.78±9.20 | 61.91±9.21 | 61.46±9.17 | 0.615* |
| Sex: |  |  |  | 0.064<sup>#</sup> |
| Male | 290 (47.7) | 803 (53.2) | 292 (47.2) |  |
| Female: | 318 (52.3) | 1056 (56.8) | 326 (52.8) |  |
| BMI (Kg/m<sup>2</sup>) | 28.18±4.78 | 28.22±4.65 | 28.13±4.53 | 0.202* |
| Total knee replacement: |  |  |  | **0.013<sup>#</sup>** |
| Yes | 50 (8.2) | 199 (10.7) | 83 (13.4) |  |
| No | 558 (91.8) | 1660 (89.3) | 535 (86.6) |  |

Values are mean±standard deviation or number of patients with percentage in parentheses; (\*) Kruskal-Wallis test; (#) chi-square test; PRS: polygenic risk score; BMI: body mass index; significant p-values are in **bold**.

To assess the incremental value of the PRS, we removed this predictor from the regression model and compared the metrics between both models (with and without PRS). The complete model reached an AUC of 0.765 (0.736 – 0.793), with a sensitivity of 0.770 and a specificity of 0.668. The clinical model (without PRS) showed a slightly lower AUC of 0.761 (0.733 – 0.789), with a sensitivity of 0.808 and a specificity of 0.604. However, despite the slight improvement in the AUC value, no significant differences were detected between the AUC of the clinical model (without PRS) and the AUC of the complete model (p=0.385). Calibration was similar for the complete and clinical models (Brier score 0.097 for both), showing broadly acceptable agreement between predicted probabilities and observed outcomes, with minor deviations at the extremes as suggested by the decile-based calibration plots (**Supplementary** Figures 1a and 1b).

In view of the positive impact that the addition of PRS has on specificity (from 0.604 to 0.668), we subsequently analyzed if the improvement provided by this score in terms of specificity was significantly different between the clinical model and the complete model. McNemar test (Bonferroni-adjusted) showed significantly improved specificity in the complete vs. clinical model at several fixed sensitivities: 0.400 (p-value=0.018), 0.450 (p-value=0.028), 0.550 (p-value=0.015), 0.700 (p-value=3.12e^-05^), and 0.750 (p-value=5.94e^-09^) (**Supplementary** Figures 2). By selecting the highest statistically significant sensitivity of 0.750 to construct the confusion matrices, the specificity increased significantly from 0.640 for the clinical model to 0.676 for the complete model (**Supplementary** Figure 3).

### KL-stratified analysis

We re-estimated the Complete model within baseline strata KL 0–1 (n=1,071; 16 TKR events/1,055 non-TKR events) and KL ≥2 (n=1,336; 275 TKR events/1,061 non-TKR events). In KL 0–1, only age (p-value=0.035) and the standardized PRS (p-value=0.002) remained significant; performance was good (AUC 0.827, 95% CI 0.715–0.940) with acceptable calibration (Brier 0.014),. In KL ≥2, several clinical predictors were significant and the PRS retained an independent association (p-value=0.028); overall performance was moderate (AUC 0.720, 95% CI 0.687–0.753; Brier 0.149). In both strata, calibration was overall adequate, with small deviations at the extremes for KL ≥2, and greater imprecision in KL 0–1 probably due to the low number of TKR events in that stratum (**Table 5** and **Supplementary** Figures 1c and 1d).

**Table 5.** Multivariate regression analyses to predict the risk of TKR KL 0-1 and KL ≥2 patients.

|  | KL 0-1 strata (n=1,071) |  | KL ≥2 strata (n=1,336) |  |
| --- | --- | --- | --- | --- |
|  | OR (95% CI) | p-value | OR (95% CI) | p-value |
| Sex (female) | 1.834 (0.563-5.975) | 0.314 | 1.045 (0.777-1.405) | 0.772 |
| Age at baseline (years) | 1.069 (1.005-1.139) | **0.035** | 1.021 (1.003-1.038) | **0.019** |
| BMI (Kg/m<sup>2</sup>) | 0.960 (0.840-1.097) | 0.546 | 1.015 (0.984-1.048) | 0.339 |
| *TP63*-GGxUK<sup>§</sup> |  |  |  |  |
| GGxUk | 1 | - | 1 | - |
| GGxno-Uk | 0.384 (0.034-4.271) | 0.436 | 0.548 (0.310-0.970) | **0.04** |
| No-GGxUk | 0.553 (0.051-6.010) | 0.627 | 0.586 (0.325-1.055) | 0.07 |
| No-GGxNo-Uk | 0.553 (0.054-4.336) | 0.519 | 0.654 (0.394-1.085) | 0.1 |
| Standardized PRS (per SD) | 2.310 (1.353-3.944) | **0.002** | 1.174 (1.017-1.355) | **0.028** |
| Systolic pressure (mm Hg) | 2.754 (0.892-8.507) | 0.078 | 1.328 (0.993-1.777) | 0.056 |
| NSAID exposure: |  |  |  |  |
| No-exposure | 1 | - | 1 | - |
| Exposure | 3.192 (0.852-11.943) | 0.085 | 1.999 (1.430-2.793) | **<0.001** |
| Previous knee injury |  |  |  |  |
| No-injury | 1 | - | 1 | - |
| Injury | 0.888 (0.300-2.623) | 0.830 | 1.118 (0.833-1.501) | 0.457 |
| Knee alignment: |  |  |  |  |
| Normal | 1 | - | 1 | - |
| Varus | 1.110 (0.288-4.269) | 0.879 | 1.478 (0.956-2.285) | 0.08 |
| Valgus | 0.550 (0.158-1.907) | 0.346 | 1.425 (0.951-2.133) | 0.08 |
| WOMAC pain | 1.091 (0.966-1.233) | 0.160 | 1.109 (1.079-1.141) | **<0.001** |
| aMED score | 0.977 (0.883-1.081) | 0.658 | 0.989 (0.960-1.019) | 0.464 |

TKR: total knee replacement; OAI: Osteoarthritis initiative; BMI: body mass index; PRS: polygenic risk score; SD: standard deviation; NSAID: non-steroid anti-inflammatory drug; WOMAC: Western Ontario and McMaster Universities Osteoarthritis Index; aMED: adherence to Mediterranean diet; OR: odds ratio; CI: confidence interval; Q1: quintile 1; (<sup>§</sup>) GG genotype of single nucleotide polymorphism (SNP) rs12107036 at *TP63* and mitochondrial DNA haplogroup Uk; (\*) significant after univariate analysis; (#) significant after multivariate analysis; significant p-values are in **bold**.

## DISCUSSION

Our primary aim was to build a pragmatic prediction model for TKR in knee OA patients from the OAI using routinely available, non-imaging variables, guided by ML feature selection. This strategy identified a compact set of clinically interpretable predictors, some of them novel (i.e MD), and delivered overall performance that meets commonly accepted clinical thresholds. Notably, calibration was broadly acceptable, supporting the model’s potential utility for risk communication and patient stratification in settings where imaging is not readily available or cost-effective.

The constructed model incorporated variables previously linked to OA and TKR. WOMAC pain and physical function, for instance, are well-established predictors [23]. A novel finding was the inverse relationship between MD adherence (aMED score) and TKR risk. Although not previously shown in this context, MD adherence has been associated with lower OA prevalence, less pain worsening, and improved cartilage quality on MRI in OAI participants [29, 34, 35]. MD’s anti-inflammatory and antioxidant properties likely mediate these protective effects [36].

Another relevant factor was systolic blood pressure. Patients with higher values had greater TKR risk. This aligns with prior evidence linking hypertension to incident knee OA, even after adjusting for BMI [37]. Similarly, other components of metabolic syndrome (MetS) and hypertension have been associated with TKR risk [38]. The effects of hypertension, as well as of other components of MetS, could result in vessel narrowing and reduced blood flow, leading to cartilage degeneration due to reduced nutrient from the subchondral bone [38]. In this sense, NSAID use, another top predictor, may increase blood pressure but may also reflect more severe disease or symptomatic burden [39, 40]. In agreement with this, prior studies have also linked prolonged NSAID use to joint replacement [41]. Interestingly, in our cohort, a higher aMED score was correlated with lower diastolic pressure (r = –0.104; p-value < 0.001), reinforcing potential dietary effects on vascular health.

We also identified a significant gene–mitochondrial DNA interaction: the GG genotype of rs12107036 in *TP63* combined with haplogroup UK was associated with increased TKR risk. This interaction was previously linked to rapid knee OA progression in the OAI cohort [28], consistent with the fact that accelerated OA increases the likelihood of needing a prosthesis [42]. Other key predictors, such as knee malalignment, prior knee injury, age, and BMI, are well-documented TKR risk factors [43, 44]. Our results reinforce their continued importance, even when genetic and nutritional data are included.

As a secondary analysis, we examined whether genetic information adds value beyond clinical predictors. To do this, we constructed a TKR-related PRS derived from one of the largest GWAS meta-analysis in OA [13], being the first study to replicate this score in the OAI cohort. The predictive performance of PRS alone was modest (AUC < 0.6) (data not shown), consistent with prior reports [17, 18]. This limited performance likely reflects the multifactorial nature of TKR risk, where clinical and lifestyle factors may overshadow genetic contributions, particularly in older individuals [45]. In this sense, in the pre-radiographic/early stratum (baseline KL 0–1), the standardized PRS remained independently associated with TKR risk and was among the very few variables maintaining statistical significance, whereas many clinical predictors weakened. This pattern is biologically plausible since genetic signal may be relatively more informative before structural disease and symptom burden fully emerge; yet it must be interpreted cautiously given the small number of events (16 TKRs) in this stratum. Interestingly, among PRS variants, rs34195470 (*WWP2*) and rs34616559 (*SOX5*) stood out. Both genes are OA-related and considered promising therapeutic targets. *WWP2* is the host gene of the microRNA-140 (*miR140*) involved in *TGF-*β signaling and cartilage maintenance [46, 47]. In addition to its association with TKR in the base GWAS used in this study [13], this SNP was also previously associated with knee OA risk in a large GWAS meta-analysis [48]. On the other hand, *SOX5* is a transcription factor crucial for cartilage development in terms of chondrogenesis and the maintenance of cartilage homeostasis [49], along with *SOX6* and *SOX9*, and its decreased expression is linked to advanced OA [50].

This study has some limitations. The lack of external validation restricts generalizability, as few cohorts combine deep genetic, nutritional, and clinical data. To strengthen internal reliability, we used a nested 5×5-fold cross-validation for feature selection, 10-fold cross-validation with SMOTE for hyperparameter tuning, and repeated 5-fold cross-validation (5 repeats) to stabilize AUC estimates, all within the training set before a final hold-out evaluation. In addition, a notable strength of our study is the dual use of linear (Elastic Net) and nonlinear (XGBoost) ML approaches, which revealed complementary patterns: Elastic Net emphasized variables with linear effects (e.g., PRS), while XGBoost captured complex, nonlinear relationships. In addition, a key strength of our work is adherence to the TRIPOD+AI reporting guidelines, which promote transparency and reproducibility in prediction model studies. While not all items are applicable or fully addressed in our context, we provide a detailed checklist in **Supplementary Table 2**.

In summary, we used ML to develop a practical, non-imaging model that predicts TKR with clinically acceptable performance. Adding the PRS modestly improved specificity, useful for reducing false positives and refining stratification, while having minimal impact on AUC, so its standalone clinical utility remains uncertain. Looking forward, as larger sequencing efforts yield more informative PRS, genetic risk could help triage early OA (KL 0–1) toward targeted MRI precisely where clinical signals are weak and imaging can be inconclusive. We deliberately excluded imaging here because MRI, though informative, is costly and not routinely available, and KL grade, often the strongest imaging-derived predictor, mainly reflects advanced disease (KL 3–4), limiting its relevance in early-stage OA (KL 0–1) [51]. A tiered pathway may enhance cost-efficiency and enable earlier intervention, as others have proposed [22, 52]. On the other hand, the results of this study also show the impact of a series of factors, some of which are either preventable (blood pressure, diet, previous injury) or indicative of personalized care for OA patients (knee alignment, PRS, genetic interaction).

## Conclusions

Our study shows that a ML–guided, non-imaging approach can produce a practical and transparent model for predicting TKR risk in knee OA. This framework advances precision medicine by turning routinely collected clinical and lifestyle data, augmented where appropriate by genetics, into individualized risk estimates that can guide prevention, targeted imaging, and referral pathways. Such models offer a scalable route to more timely, patient-specific decisions. Next steps are rigorous external validation and implementation research to ensure reliable performance and equitable impact across settings.

## Data Availability

Data are available upon reasonable request

## Abbreviations

AUC: Area Under the Curve
aMED: Alternate Mediterranean Diet Score
BMI: Body Mass Index
CI: Confidence Interval
dbGAP: Database of Genotypes and Phenotypes
GBM: Gradient Boosting Machine
GeCKO: Genetic Components of Knee Osteoarthritis
glmnet: Generalized Linear Model Net
GWAS: Genome-Wide Association Study
KL: Kellgren & Lawrence
MAF: Minor Allele Frequency
MD: Mediterranean Diet
ML: Machine Learning
MRI: Magnetic Resonance Imaging
mtDNA: Mitochondrial Deoxyribonucleic Acid
NSAIDs: Non-Steroidal Anti-Inflammatory Drugs
OA: Osteoarthritis
OAI: Osteoarthritis Initiative
OR: Odds Ratio
PCA: Principal Component Analysis
PRS: Polygenic Risk Score
QC: Quality Control
RF: Random Forest
ROC: Receiver Operating Characteristic
SHAP: SHapley Additive exPlanations
SNP: Single Nucleotide Polymorphism
SMOTE: Synthetic Minority Oversampling Technique
SVM: Support Vector Machine
TKR: Total Knee Replacement
VCF: Variant Call Format
WOMAC: Western Ontario and McMaster Universities Osteoarthritis Index
XGBoost: Extreme Gradient Boosting

## Acknowledgements

The Osteoarthritis Initiative (OAI) is a public-private partnership comprised of five contracts (N01-AR-2-2258; N01-AR-2-2259; N01-AR-2-2260; N01-AR-2-2261; N01-AR-2-2262) funded by the National Institutes of Health, a branch of the Department of Health and Human Services, and conducted by the OAI Study Investigators. Private funding partners include Pfizer, Inc.; Novartis Pharmaceuticals Corporation; Merck Research Laboratories; and GlaxoSmithKline. Private sector funding for the OAI is managed by the Foundation for the National Institutes of Health. This manuscript was prepared using an OAI public use data set and does not necessarily reflect the opinions or views of the OAI investigators, the NIH, or the private funding partners.

## Authoŕs contribution

FJB and IRP contributed equally in the design and coordination of the study; both conceived the study, participated in its design and helped to draft the final version of the manuscript; NO coordinated the access to the biological samples and the clinical information used in this study; VBB and IRV contributed equally in the statistical analysis of data; NV and MCA helped provide the data on the Mediterranean diet; JVG and AMT helped in the analysis of the GWAS data for PRS calculation; SR performed the genetic assignments of mtDNA haplogroups and the *TP63* genotypes.

## Ethical approval information

This study has received the favorable opinion of the ethics committee of XUNTA de Galicia, registration number 2024/074

## Data sharing statement

Data are available upon reasonable request

## Patient and Public Involvement

To the best of our knowledge, patients OAI cohort are informed about the goal of their enrollment in these prospective studies. The entity responsible of this cohort will be the first to receive a copy of this manuscript once it is published. In addition, all the clinical centers involved in the recruitment of the patients have made provisions to ensure the safety, confidentiality and ethical treatment of study participants according to the Declaration of Helsinki. In this sense, all the participants signed an informed consent

## Declaration of generative AI and AI-assisted technologies in the writing process

During the preparation of this work the author(s) used ChatGPT in order to adjust the different R scripts. After using this tool, the author(s) reviewed and edited the content as needed and take(s) full responsibility for the content of the publication.

---

**Supplementary Figure 1.** Decil-based calibration plots to compare the models. Predictions were split into ten equal groups; for each, we plotted the average predicted probability against the observed event rate, adding binomial-test confidence bounds and the 45° reference line. **A)** Complete model; **B)** Complete model without PRS; **C)** Complete model for KL 0-1 stratum; **D)** Complete model for KL ≥2 stratum.

**Supplementary Figure 2.** p-values of the McNemar test to compare the specificity values between the clinical model and the complete model with PRS.

**Supplementary Figure 3.** Confusion matrices for a fixed sensitivity of 0.75 for the clinical model and the complete model with PRS.

