## Supplementary figures and images for "Predicting Total Knee Replacement in Knee Osteoarthritis Using a Machine-Learning–Guided Approach in patients of the Osteoarthritis Initiative (OAI)"

### Supplementary Figure 1a

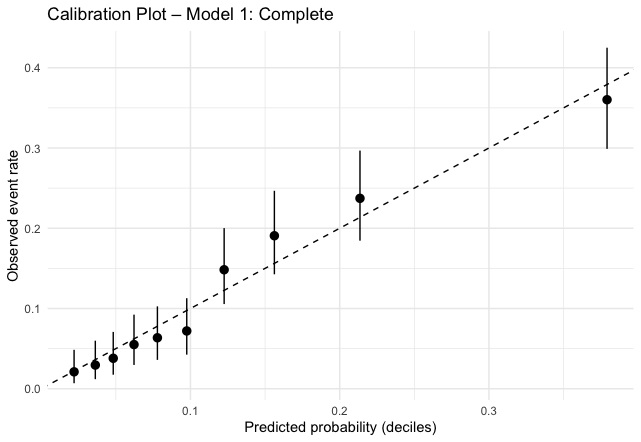

### Supplementary Figure 1b

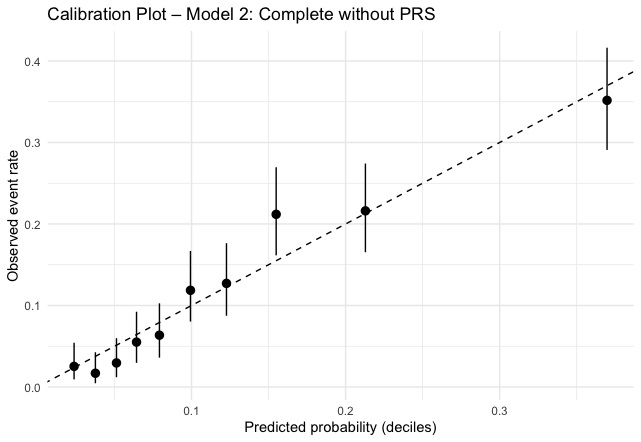

### Supplementary Figure 1c

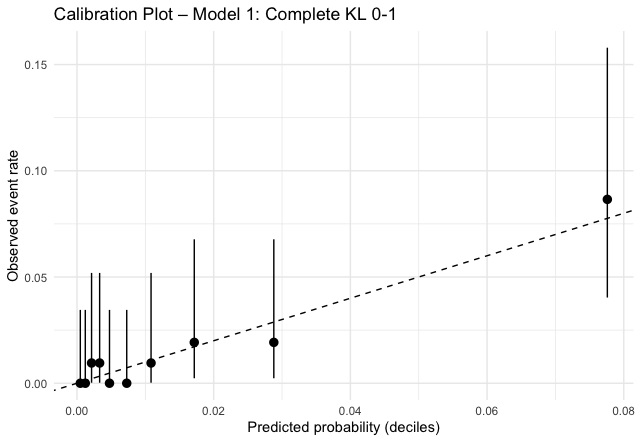

### Supplementary Figure 1d

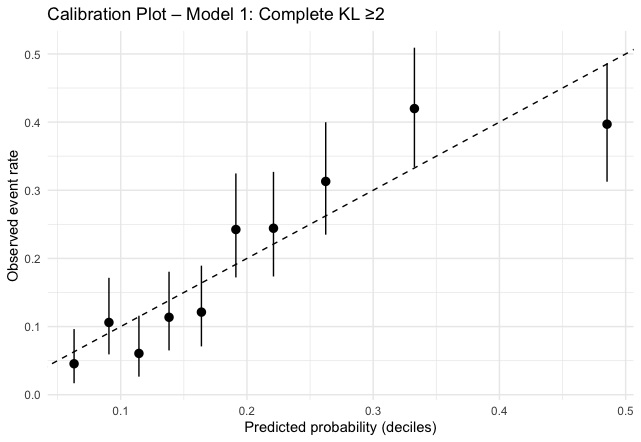

### Supplementary Figure 2

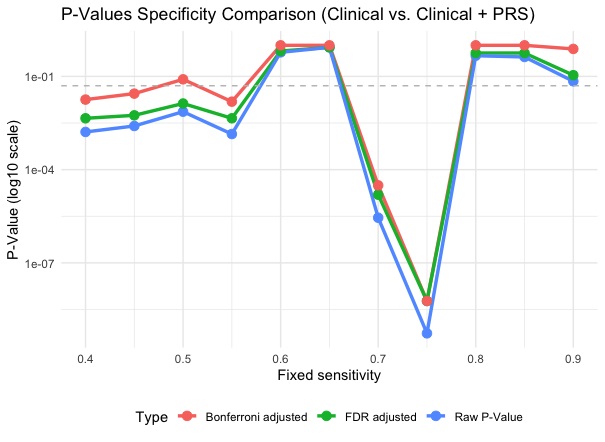

### Supplementary Figure 3

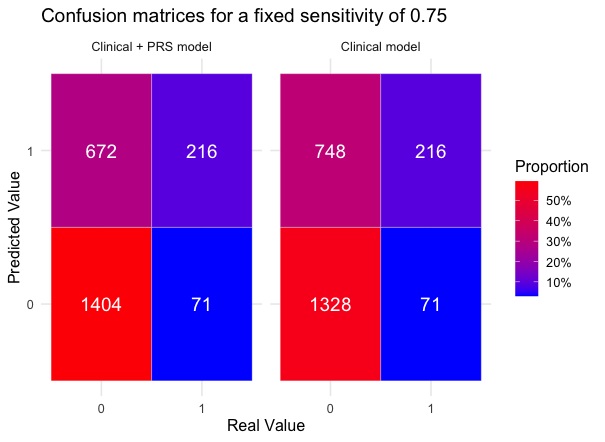
